# Cardiomyopathy in patients with acute ischemic stroke and methamphetamine use: Relevance for cardioembolic stroke and outcome

**DOI:** 10.1101/2023.11.22.23298930

**Authors:** Sook Joung Lee, Shimeng Liu, Haley Blackwill, Dana Stradling, Mohammad Shafie, Wengui Yu

## Abstract

Methamphetamine use has emerged as a major risk factor of cardiovascular and cerebrovascular disease in young adults. The aim of this study was to investigate the relevance of methamphetamine use and cardiomyopathy in patients with acute ischemic stroke.

We performed a retrospective study of consecutive patients with acute ischemic stroke admitted at our medical center between 2019 and 2022. All patients were screened for methamphetamine use and cardiomyopathy defined as left ventricular ejection fraction ≤ 45%. Methamphetamine use was identified by self-reported history of use and/or positive urine drug screen. Logistic regression model was performed to analyze the relevance of methamphetamine-associated cardiomyopathy and cardioembolic stroke.

Among 973 consecutive patients screened for the study, 48 (4.9%) were identified to have methamphetamine use. Compared with Non-meth group (n=892), the patients in the Meth group were significantly younger (53.2 ± 10.0 vs. 69.7 ± 15.2, p <0.001), more likely male (76.6% vs. 50.0%, p <0.001), and associated with significantly higher rate of cardiomyopathy (30.4% vs. 14.0%, p <0.01). They were also less likely to have history of atrial fibrillation (8.7% vs. 33.4%, p<0.01) or hyperlipidemia (28.3% vs. 51.7%, p <0.01). Compared with patients with cardiomyopathy without methamphetamine use, those with methamphetamine use and cardiomyopathy had better functional outcome at 3 months, likely due to younger age and fewer comorbidities. In the logistic regression model, methamphetamine-associated cardiomyopathy was found to be a significant risk factor of cardioembolic stroke (OR 2.88, 95% CI 1.81-4.58, p < 0.0001).

Our results demonstrate that methamphetamine use increases the risk of cardiomyopathy and cardioembolic stroke in young adults.

## INTRODUCTION

Methamphetamine is a synthetic psychostimulant that is highly addictive [1-2]. Methamphetamine use has emerged as a major risk factor for acute ischemic stroke (AIS) in young adults worldwide in recent years [3-12].

The mechanisms by which methamphetamine causes acute ischemic stroke are unclear. Case studies and forensic analysis showed cerebral vasoconstriction, atherosclerotic stenoses, arterial dissection, vasculitis, and small vessel disease in patients with methamphetamine-associated stroke [2,6,8,12,13]. Methamphetamine use was also shown to produce a dose-dependent elevation of blood pressure and chronic hypertension [14-15]. Hypertension, vasoconstriction, and vascular toxicity were postulated as major mechanisms of ischemic stroke [13].

Methamphetamine use also induces sympathetic activation, cardiovascular injury, dilated cardiomyopathy (CMP) with significantly reduced left ventricle ejection fraction (EF) [16-17]. Compared with non-abusers, methamphetamine abusers have more severe dilated CMP on echocardiography [18]. Methamphetamine use has been increasingly identified as a major cause of cardiomyopathy and cardiac death in young adults [19-21].

There were isolated case reports on methamphetamine-associated cardiomyopathy and cardioembolic stroke [22, 23]. In a case series of methamphetamine-associated cardiomyopathy, one-third of the patients were found to have intraventricular thrombi on endomyocardial biopsy [19], suggesting a possible mechanism of cardioembolic stroke.

The aim of this study was to investigate the prevalence of cardiomyopathy in patients with acute ischemic stroke and methamphetamine use, and its relevance to cardioembolic stroke and outcome.

## METHODS

This is a retrospective cohort study. The study protocol was approved by the University of California Irvine Institutional Review Board (IRB) and the Ethics Committee. Informed consents were waived due to retrospective study design and minimal harm to the patients. All methods in the study were performed in accordance with the relevant guidelines and regulations.

### Study population

Consecutive AIS patients admitted at the University of California Irvine Medical Center from January 1, 2019 to December 30, 2022 were screened for the study. The patient list was generated by searching the Vizient Clinical Database using the International Classification of Diseases, 10th revision (ICD-10) codes for ischemic stroke or cerebral infarction as a primary or secondary discharge diagnosis. Vizient contains data from over 97% of US academic medical centers, including ours [24]. The following information was collected from the Vizient database and independent chart review from our electronic medical record system EPIC: age, gender, race, past medical history, home medications, social history including smoking and recreational drug use, National Institutes of Health Stroke Scale (NIHSS) scores, urine drug screen (UDS) results, LDL cholesterol levels, echocardiogram results, length of stay (LOS) in the intensive care unit and hospital, and functional outcome with modified Rankin Scale (mRS) scores at 3 months. All patients underwent standard diagnostic evaluation and treatment per American Heart Association (AHA)/American Stroke Association (ASA) guidelines [25].

Based on the history of methamphetamine abuse and urine drug screen (UDS), patients were divided into Meth and Non-meth group. The UDS was performed using EMIT II Plus Amphetamines Assay (Beckman Coulter, Inc) with a sensitivity and specificity of 94.3% and 93.3%, respectively [26]. Cardiomyopathy was defined with reduced left ventricular ejection fraction to 45% or less on clinical echocardiography report [18].

### Statistical analysis

Continuous variables were described by mean ± standard deviation (SD) on the results of normality testing. Categorical variables were expressed by counts with percentages. Baseline characteristics and functional outcome at 3 months as determined by mRS were compared between Meth and Non-meth groups by the student t-test for continuous variables and chi-square test for categorical variables. Logistic regression model was performed to identify independent predictors of cardioembolic stroke. All statistical analyses were performed using SAS software. A 2-tailed value of *p* < 0.05 was considered statistically significant.

## RESULTS

973 consecutive patients were admitted for AIS during the study period (Figure 1). Thirty patients were excluded from the study due to incomplete data, lack of echocardiogram, or lost to follow-up. Among the remaining patients, 48 (4.9%) were identified to have a history of methamphetamine use or positive UDS. Two patients in the Meth group and 3 patients in the Non-meth group had concomitant cocaine use and were therefore excluded from the study.

**Figure 1.**
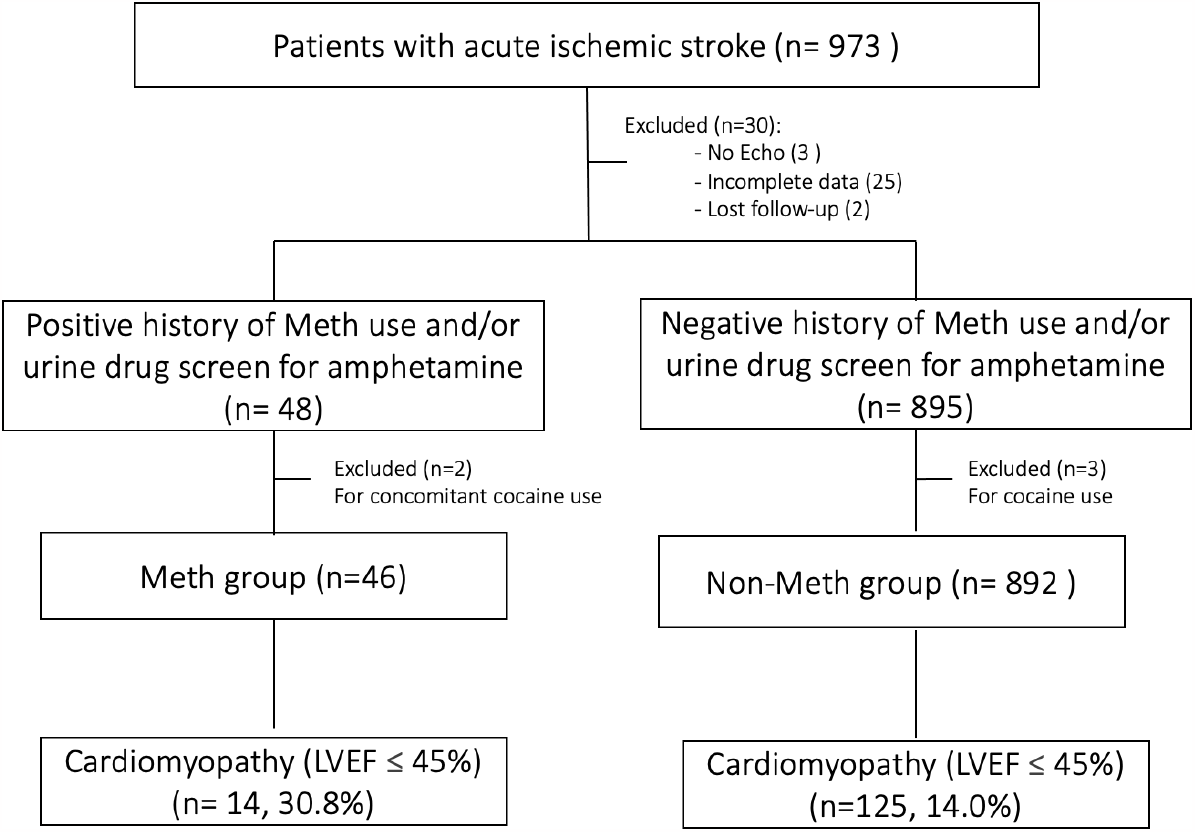
The Study flowchart.

Among the 46 remaining patients in the Meth group, 14 (30.4%) were found to have cardiomyopathy (CMP) on clinical echocardiogram report. In contrast, 125 patients (14%) in the Non-meth group were noted to have cardiomyopathy.

The basic characteristics of the Meth and Non-meth groups were shown in Table 1. Compared with the Non-meth group (n=892), the patients in the Meth group were significantly younger (53.2 ± 10.0 vs. 69.7 ± 15.2, p <0.001), more likely male (76.6% vs. 50.0%, p <0.001) and White (43.5% vs. 30.5%, p <0.05), and less likely Asians (13% vs.27.5%, p <0.05). They were less likely to have history of atrial fibrillation (8.7% vs. 33.4%, p <0.01), hyperlipidemia (28.3% vs. 51.7%, p <0.01), or statin use (30.4% vs. 47.0%, p <0.05, but more likely to have history of smoking (95.7% vs. 24.8%, p <0.001), heart failure (37.0% vs. 22.9%, p <0.05), cardiomyopathy (30.4% vs. 14.0%, p <0.01), or small vessel disease as etiology of stroke (25.5% vs 10.3%, p <0.01). There were no significant differences between the 2 groups in history of hypertension, diabetes, obesity, the use of antiplatelet or anticoagulation medications, initial NIHSS scores, LDL cholesterol levels, echocardiogram findings of patent foramen ovale or left ventricular thrombus, cardioembolism as cause of stroke, LOS in the ICU or hospital, good functional outcome (mRS 0-2) and mortality at 3 months.

**Table 1.** Demographics and clinical Characteristics between Meth group and Non-meth group.

| Variables | Meth group | Non-meth group | P value |
| --- | --- | --- | --- |
| N | 46 | 892 |  |
| Age | 52.8 ± 9.64 | 69.7 ± 15.2 | < 0.001 |
| Female (%) | 10 (21.7%) | 421 (50.0%) | < 0.001 |
| Race |  |  |  |
| White | 20 (43.5%) | 272 (30.5%) | < 0.05 |
| Hispanic | 8 (17.4%) | 131 (14.7%) | 0.66 |
| Black | 8 (17.4%) | 197 (20.1%) | 0.41 |
| Asian | 6 (13.0%) | 243 (27.2%) | < 0.05 |
| Other | 4 (8.7%) | 49 (5.5%) | 0.38 |
| Hypertension | 42 (91.3%) | 771 (86.4%) | 0.32 |
| Diabetes | 18 (39.1%) | 400 (44.8%) | 0.78 |
| Hyperlipidemia | 13 (28.3%) | 461 (51.7%) | < 0.01 |
| Atrial fibrillation | 4 (8.7%) | 300 (33.4%) | < 0.01 |
| Heart failure | 17 (37.0%) | 204 (22.9%) | < 0.05 |
| Obesity (BMI >30) | 22 (47.8%) | 317 (35.5%) | 0.12 |
| Smoking | 44 (95.7%) | 221 (24.8%) | < 0.001 |
| Antiplatelet | 11 (23.9%) | 260 (28.1%) | 0.5415 |
| Anticoagulant | 2 (4.4%) | 134 (14.5%) | 0.0536 |
| Statin use | 14 (30.4%) | 436 (47.0%) | < 0.05 |
| Initial NIHSS score | 8.61 ± 8.00 | 10.32 ± 8.64 | 0.29 |
| LDL cholesterol | 85.36 ± 30.62 | 89.62 ± 41.85 | 0.37 |
| Echo finding |  |  |  |
| LV EF (≤45%) | 14 (30.4%) | 125 (14.0%) | < 0.01 |
| Positive PFO (n, %) | 3 (6.5%) | 81 (9.1%) | 0.53 |
| LV Thrombus | 4 (8.7%) | 78 (8.7%) | 0.96 |
| Mechanism of stroke |  |  |  |
| Large artery disease | 9 (19.6%) | 290 (32.5%) | 0.55 |
| Cardioembolism | 20 (43.5%) | 408 (45.7%) | 0.89 |
| Small vessel disease | 12 (26.1%) | 92 (10.3%) | < 0.01 |
| Other causes | 5 (10.9%) | 125 (14.0%) | 0.51 |
| ICU LOS, median (IQR) | 3.0 (2.0-7.0) | 3.0 (1.0-5.0) | 0.5271 |
| Hospital LOS, median (IQR) | 4.5 (2.0-8.0) | 4.0 (2.0-7.0) | 0.4179 |
| mRS 0-2 at 3-month | 28 (60.9%) | 517 (58.0%) | 0.83 |
| Mortality at 3-month | 4 (8.7%) | 171 (19.2%) | 0.14 |

LDL, low density lipoprotein cholesterol; mRS, modified Rankin score; NIHSS, National Institutes of Health Stroke Scale; PFO, patent foramen ovale.

**Table 2.** Characteristics of cardiomyopathy patients with and without methamphetamine use

| Variables | Meth CMP group | Non-Meth CMP group | P value |
| --- | --- | --- | --- |
| N | 14 | 125 |  |
| Age | 52.4 ± 9.5 | 70.9 ± 15.4 | < 0.001 |
| Female (%) | 3 (21.4%) | 46 (36.8%) | 0.25 |
| Race |  |  |  |
| White | 5 (35.7%) | 43 (34.4%) | 0.92 |
| Hispanic | 4 (28.6%) | 24 (19.2%) | 0.41 |
| Black | 1 (7.1%) | 19 (15.2%) | 0.42 |
| Asian | 2 (14.3%) | 30 (24.0%) | 0.41 |
| Other | 2 (14.3%) | 9 (7.2%) | 0.35 |
| Hypertension | 12 (85.7%) | 124 (99.2%) | < 0.01 |
| Diabetes | 5 (35.7%) | 54 (43.2%) | 0.59 |
| Hyperlipidemia | 3 (21.4%) | 62 (49.6%) | < 0.05 |
| Atrial fibrillation | 2 (14.3%) | 62 (49.6%) | < 0.05 |
| Heart failure | 12 (85.7%) | 98 (78.4%) | 0.52 |
| Obesity (BMI >30) | 6 (42.9%) | 46 (36.8%) | 0.66 |
| Smoking | 14 (100%) | 29 (23.2%) | < 0.001 |
| Initial NIHSS score | 10.71 ± 8.18 | 13.80 ± 8.59 | 0.10 |
| LDL cholesterol | 77.85 ± 23.66 | 81.90 ± 45.29 | 0.38 |
| Echo finding |  |  |  |
| LV EF | 26.0 ± 9.59 | 32.47 ± 9.52 | < 0.01 |
| Positive PFO (n, %) | 2 (14.3%) | 8 (6.4%) | 0.28 |
| LV Thrombus | 4 (28.6%) | 24 (19.2%) | 0.41 |
| Mechanism of stroke |  |  |  |
| Large artery disease | 0 (0.0%) | 23 (18.4%) | 0.29 |
| Cardioembolism | 13 (92.9%) | 91 (72.8%) | 0.10 |
| Small vessel disease | 1 (7.1%) | 5 (4.0%) | 0.58 |
| Other causes | 1 (7.1%) | 7 (5.6%) | 0.81 |
| mRS 0-2 at 3-month | 9 (64.3%) | 44 (35.2%) | < 0.05 |
| Mortality at 3-month | 2 (14.3%) | 43 (34.4%) | 0.13 |

LDL, low density lipoprotein cholesterol; mRS, modified Rankin score; NIHSS, National Institutes of Health Stroke Scale. P value < 0.05 was considered statistically significant.

In subgroup analysis of cardiomyopathy patients with and without methamphetamine use (Table in 2), the patients with methamphetamine use (Meth CMP group, n =14) remained significantly younger (52.4 ± 9.5 vs. 70.9 ± 15.4, p <0.001) and more likely smoker (100% vs. 23.3%, p<0.001) than those without methamphetamine use (Non-meth CMP group, n=125). The patients with methamphetamine use were also found to have significantly lower left ventricle ejection fraction (26.0 ± 9.59 vs. 32.47 ± 9.52, p <0.01) than those without methamphetamine use. In contrast, patients in the Non-meth-CMP group had significantly higher rates of hypertension, hyperlipidemia, and atrial fibrillation than those in the Meth CMP group.

Of note, despite significantly lower left ventricle ejection fraction, the patients in the Meth CMP group had significantly higher rate of favorable functional outcomes (mRS 0-2) at 3-month after AIS than the Non-meth CMP group (64.3% vs. 35.2%, p <0.05).

Logistic regression model was performed for multivariate analysis of risk factors of cardioembolic stroke. As shown in Table 3, among all clinical variables, only 2 were identified to be significantly associated with cardioembolic stroke. Atrial fibrillation was the most significant risk factor for cardioembolic stroke in the entire cohort (OR 20.89, 95% CI 14.112-30.91, p <0.0001). This was followed by methamphetamine-associated cardiomyopathy (OR 2.88, 95% CI 1.81-4.58, p <0.0001).

**Table 3.** Positive predictors of cardioembolic stroke in patients with acute ischemic stroke.

|  | OR of cardioembolic stroke | 95% CI | P |
| --- | --- | --- | --- |
| A Fib | 20.89 | 14.12 - 30.91 | < 0.0001 |
| Meth-associated CMP | 2.88 | 1.81 – 4.58 | <0.0001 |

A Fib, atrial fibrillation; CMP, cardiomyopathy; OR, odds ratios.

## DISCUSSION

Our results demonstrate that methamphetamine abuse was seen in 4.9% of the patients with AIS at our medical centers. Compared with AIS patients without methamphetamine use, patients with methamphetamine use were significantly younger and more likely male. They were less likely to have atrial fibrillation but highly associated with cardiomyopathy and propensity for cardioembolic stroke. Of note, the patients with methamphetamine-associated cardiomyopathy had better functional outcome at 3-month after stroke than those without methamphetamine use, possibly due to younger age and fewer comorbidities such as atrial fibrillation and hyperlipidemia.

Our cohort study corroborates previous reports on the younger and male predominance in patients with methamphetamine-associated cardiomyopathy [10, 27-29] and stroke [11, 12] . Public education and government policies are needed to increase the awareness of the major health concerns of methamphetamine use and the socioeconomic burden of methamphetamine associated disabilities [20,21].

The mechanisms of methamphetamine associated cardioembolic stroke remain unclear but are likely multifactorial. Methamphetamine may cause vasoconstriction, atherosclerotic disease, cardiac arrhythmias, and cardiomyopathy [30]. It was also shown to promote myocardial structural or electrical remodeling, such as increased ventricular fibrosis, inflammation, or myocyte function [17, 31,32]. These remodeling of cardiac tissue following methamphetamine exposure promotes dilated cardiomyopathy and the susceptibility to cardiac arrhythmia and heart failure [17,30]. It has been theorized that heart failure, a clinically significant reduction in left ventricle EF associated with progressive LV dilatation and cardiac remodeling, will have a heighted risk for cardioembolic stroke [33]. Our study corroborated findings from previous reports that methamphetamine abusers have significantly lower left ventricle EF than non-abusers [17, 20, 28,29]. Of note, methamphetamine associated non-ischemic cardiomyopathy can be reversible and that methamphetamine cessation is associated with improvement in functional status [10, 23, 33].

Currently, there were only isolated case reports suggesting the association between cardiomyopathy and cardioembolic stroke in patients with methamphetamine use [22, 23]. We have previously reported on methamphetamine abuse increasing the risk of cerebral small vessel disease in young patients with AIS [12]. That study also found a high percentage (34%) of cardioembolic stroke in patients with methamphetamine abuse. This cohort study demonstrated that 92.9% of patients with methamphetamine-associated cardiomyopathy had cardioembolic stroke, suggesting cardiomyopathy as the possible mechanism of cardioembolic stroke. To our knowledge, we are the first cohort study to demonstrate methamphetamine-associated cardiomyopathy and the propensity of cardioembolic stroke.

Our study has a few limitations. First, it is a retrospective study with a relatively small sample size of ethnically diverse patient population in Southern California. The results may not be generalizable. Second, our data only suggested a possible association between methamphetamine-associated cardiomyopathy and cardioembolic stroke. Third, there was no information regarding the route, frequency, and duration of methamphetamine abuse. Lastly, alcohol use and other cardiac co-morbidities could be confounding factors that cannot be ruled out in this retrospective study. Further large sample size studies are warranted to adjust for confounding factors and to establish the temporal relationship between methamphetamine-associated cardiomyopathy and cardioembolic stroke.

## CONCLUSION

In conclusion, our cohort study demonstrates that methamphetamine use increases the risk of cardiomyopathy and cardioembolic stroke in young adults.

## Data Availability

Data of this study are available from the corresponding author on reasonable request.

## Acknowledgements

Dr. Sook Joung Lee is supported by Republic of Korea for this research project.

## Authors Contributions

SJL contributed to data acquisition, statistical analysis, data interpretation and drafting manuscript.

SL contributed statistical analysis and verification.

HB and DS contributed to data acquisition.

MS contributed to data interpretation and manuscript revision.

WY contributed to study design, data interpretation, drafting and finalizing the manuscript.

## Data availability Statement

Data of this study are available from the corresponding author on reasonable request.

## Conflict of Interest Statement

All authors had no conflict of interest.

## Notes

### Competing Interest Statement

The authors have declared no competing interest.

### Clinical Trial

N/A

### Funding Statement

No external funding was received for this research project

### Author Declarations

The study protocol was approved by the University of California Irvine Institutional Review Board (IRB) and the Ethics Committee. Informed consents were waived due to retrospective study design and minimal harm to the patients.

